# Retrospective Assessment of Deep Neural Networks for Skin Tumor Diagnosis

**DOI:** 10.1101/2019.12.12.19014647

**Authors:** Seung Seog Han, Ik Jun Moon, Jung-Im Na, Myoung Shin Kim, Gyeong Hun Park, Seong Hwan Kim, Kiwon Kim, Ju Hee Lee, Sung Eun Chang

## Abstract

**BACKGROUND:** The aim of this study was to validate the performance of algorithm (http://rcnn.modelderm.com) for the diagnosis of benign and malignant skin tumors.

**METHODS:** With external validation dataset (43 disorders; 40,331 clinical images from 10,426 patients; January 1, 2008 – March 31, 2019), we compared the prediction of algorithm with the clinical diagnosis of 65 attending physicians at the time of biopsy request.

**RESULTS:** For binary-task classification of determining malignancy, the AUC of the algorithm was 0.863(95% CI 0.852-0.875) with unprocessed clinical photographs. The sensitivity/specificity of the algorithm at the predefined high-sensitivity and high-specificity threshold were 79.1%(76.9-81.4)/76.9%(76.1-77.8) and 62.7%(59.9- 65.5)/90.0%(89.4-90.6), respectively. The sensitivity/specificity calculated by the clinical diagnosis of attending physicians were 88.1%/83.8%(Top-3) and 70.2%/95.6%(Top-1), which were superior to those of algorithm.

For multi-task classification, the mean Top-1,2,3 accuracies of the algorithm were 42.6±20.7%, 56.1±22.8%, 61.9±22.9%, and those of clinical diagnosis were 65.4±17.7%, 73.9±16.6%, 74.7±16.6%, respectively.

In the reader test with images from 30-patients batches, the sensitivity / specificity of the algorithm at the predefined threshold were 66.9%±30.2% / 87.4%±16.5%. The sensitivity / specificity derived from the first diagnosis of 44 the participants were 65.8%±33.3% / 85.7%±11.0%, which were comparable with those of the algorithm (Wilcoxon signed-rank test; P=0.61 / 0.097).

**CONCLUSIONS:** Our algorithm could diagnose skin tumors at dermatologist-level when diagnosis was made solely with photographs, demonstrating its potential as a mass screening tool in telemedicine setting. However, due to limited data relevancy, the performance was inferior to that of actual medical examination. Clinical information should be integrated with imaging information to achieve more accurate predictions.

## Introduction

Deep learning algorithm has shown excellent results in analyzing fundoscopy images in ophthalmology^1^, chest X-ray^2^ and CT images in radiology.^3^ In dermatology, there have been remarkable advances that showed results comparable to dermatologists’ diagnostic performance using both clinical photography^4-9^ and dermoscopy images.^4,7,10-13^

However, it is unclear that these excellent results can be extrapolated in clinical setting as like product in other field^14^. On perspective of data relevancy, DeepMind’s AlphaGo^15^ was trained with Go data which had 100% data relevancy, and surpassed human champion. In speech recognition, gestures may be helpful in understanding conversation completely, but audio data shows data relevancy close to 100%. However, in medical field, clinical data used in training has relatively low data relevancy. For example, even if algorithm will be trained with countless brain MRIs, we cannot clearly answer whether the algorithm can truly diagnose Parkinson’s disease with MRI alone. Although image of skin contains important relevant information, there is no study of how much important in skin cancer diagnosis. This study was designed to compare the performance of algorithm with that of attending physician in real practice for the diagnosis of benign and malignant tumors.

In addition, deep learning algorithm can produce reliable result only for the preselected diseases because algorithm may show epistemic uncertainty for untrained problems^16^. Because most of previous studies had a limitation that small number of disorders were validated^17^, we investigated the generalizability of our algorithm with almost all sorts of skin tumors.

## Methods

With approvals of institutional review boards (Severance: #2019-0571 and Asan: #2017-0087), we gathered clinical photographs from Severance Hospital, Department of Dermatology, collected from January 1, 2008 to March 31, 2019. Attending physicians had recorded the clinical diagnosis on the biopsy request form considering patient’s history and physical examinations. All skin lesions of patients over 19 years of age who were pathologically diagnosed among 43 primary skin tumors were included (Figure 1). We included cases with single lesion because of the possible mismatch between the lesion and the diagnosis for patients with multiple lesions. We excluded the images of intraocular, postoperative, laser surgery, and other chief complaints, but we did not exclude normal images taken for record taking even if it did not include the lesion of interest. There were 66 cases with inadequate quality and 769 cases with which it was inherently impossible to detect lesions (Figure 1). As a result, a total of 40,331 clinical images from 10,426 patients were included in this study (Table 1).

**Figure 1.**
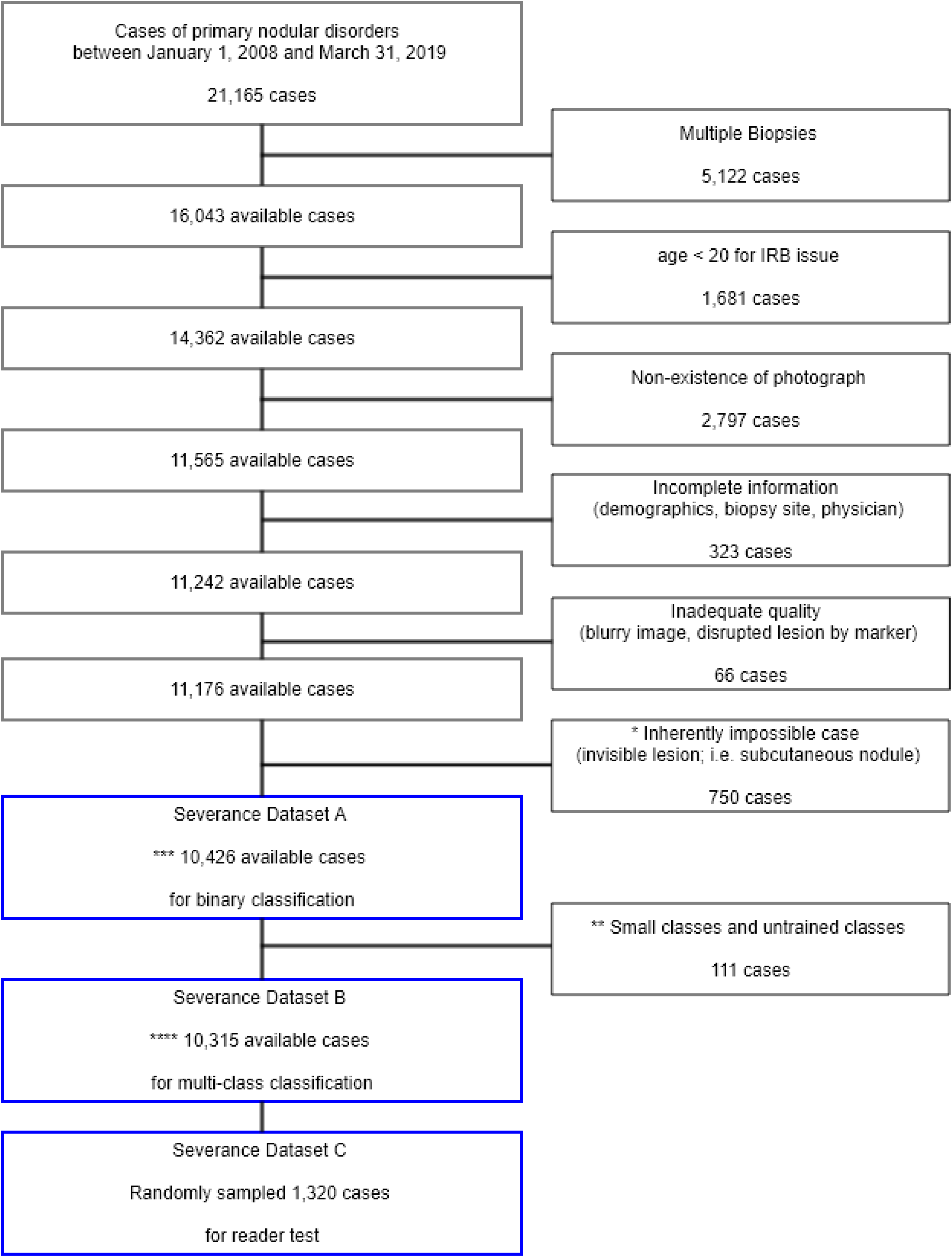
Dataset Selection Process – Exclusion Criteria. *The three most common disorders among the inherent impossible cases were epidermal cyst (497 cases), seborrheic keratosis (64 cases), and hemangioma (37 cases). There were 26 cases of malignant nodules (18 cases of basal cell carcinoma and 8 cases of other malignancies) among the inherent impossible cases. ** Small classes indicates the disease classes which has less than 10 cases: angiofibroma, Café au lait macule, juvenile xanthogranuloma, milia, nevus spilus, and sebaceous hyperplasia. Untrained classes indicates the classes which were not trained by the algorithm: dermatofibrosarcoma protuberans, Spitz nevus, Kaposi sarcoma, angiosarcoma, and Merkel cell carcinoma *** Severance Dataset A; a total of 10,426 patients (40,331 images; 43 disorders; age 52.1±18.3, male 45.1%) were used for the binary-class (cancer or not) classification. **** Severance Dataset B; a total of 10,315 patients (39,721 images; 32 disorders; age 52.1±18.2, male 44.8%) were used for the multi-class classification in the supplementary result. Among 39,721 images, a total of 34,672 images contained lesion of interests, while the remaining 5,049 images were photographs without lesion, but these photographs were taken for the purpose of observation or comparison.

**Table 1.** Demographics of the Severance Validation Dataset.

|  | Severance Dataset |  |  |
| --- | --- | --- | --- |
|  | Dataset A | Dataset B | Dataset C |
| No. of images | 40,331 | 39,721 | 5,065 |
| Patient demographics |  |  |  |
| No. of Patients | 10,426 | 10,315 | 1,320 |
| Age (mean ± SD) | 52.1 ± 18.3 | 52.1 ± 18.2 | 52.0 ± 18.3 |
| Male | 4,701 (45.1%) | 4,626 (44.8%) | 580 (43.9%) |
| No. of disorders | 43 | 32 | 31 |
| Malignancy | 1,222 (11.7%) | 1,154 (11.2%) | 152 (11.5%) |
| Angiosarcoma | 17 (0.2%) | - | - |
| Basal cell carcinoma | 643 (6.3%) | 643 (6.2%) | 101 (7.7%) |
| Dermatofibrosarcoma protuberance | 9 (0.1%) | - | - |
| Intraepithelial carcinoma (SCC in situ) | 255 (2.5%) | 255 (2.5%) | 31 (2.3%) |
| Kaposi sarcoma | 39 (0.4%) | - | - |
| Keratoacanthoma | 15 (0.1%) | 15 (0.1%) | - |
| Malignant melanoma | 83 (0.8%) | 83 (0.8%) | 13 (1.0%) |
| Merkel cell carcinoma | 3 (0.0%) | - | - |
| Squamous cell carcinoma | 158 (1.5%) | 158 (1.5%) | 7 (0.5%) |
| Benign | 9,204 (88.3%) | 9,161 (88.8%) | 1,168 (88.5%) |
| Actinic keratosis | 784 (7.6%) | 784 (7.6%) | 102 (7.7%) |
| Angiofibroma | 4 (0.0%) | - | - |
| Angiokeratoma | 39 (0.4%) | 39 (0.4%) | 4 (0.3%) |
| Becker nevus | 14 (0.1%) | 14 (0.1%) | 2 (0.2%) |
| Blue nevus | 115 (1.1%) | 115 (1.1%) | 17 (1.3%) |
| Café au lait macule | 1 (0.0%) | - | - |
| Congenital nevus | 47 (0.5%) | 47 (0.5%) | 9 (0.7%) |
| Dermatofibroma | 845 (8.2%) | 845 (8.2%) | 102 (7.7%) |
| Epidermal cyst | 1,501 (14.6%) | 1,501 (14.6%) | 203 (15.4%) |
| Epidermal nevus | 26 (0.3%) | 26 (0.3%) | 5 (0.4%) |
| Hemangioma | 263 (2.6%) | 263 (2.5%) | 34 (2.6%) |
| Juvenile xanthogranuloma | 4 (0.0%) | - | - |
| Lentigo | 67 (0.7%) | 67 (0.6%) | 2 (0.2%) |
| Lymphangioma | 15 (0.1%) | 15 (0.1%) | 1 (0.1%) |
| Melanocytic nevus | 1,441 (14.0%) | 1,441 (14.0%) | 171 (13.0%) |
| Milia | 8 (0.1%) | - | - |
| Mucocele | 73 (0.7%) | 73 (0.7%) | 10 (0.8%) |
| Mucosal melanotic macule | 36 (0.4%) | 36 (0.3%) | 7 (0.5%) |
| Neurofibroma | 199 (1.9%) | 199 (1.9%) | 31 (2.3%) |
| Nevus spilus | 3 (0.0%) | - | - |
| Organoid nevus | 62 (0.6%) | 62 (0.6%) | 7 (0.5%) |
| Ota nevus | 24 (0.2%) | 24 (0.2%) | 4 (0.3%) |
| Porokeratosis | 71 (0.7%) | 71 (0.7%) | 14 (1.1%) |
| Poroma | 64 (0.6%) | 64 (0.6%) | 10 (0.8%) |
| Portwine stain | 15 (0.1%) | 15 (0.1%) | 2 (0.2%) |
| Pyogenic granuloma | 162 (1.6%) | 162 (1.6%) | 18 (1.4%) |
| Sebaceous hyperplasia | 6 (0.1%) | - | - |
| Seborrheic keratosis | 2,370 (23.1%) | 2,370 (23.0%) | 291 (22.0%) |
| Skin tag | 70 (0.7%) | 70 (0.7%) | 8 (0.6%) |
| Spitz nevus | 17 (0.2%) | - | - |
| Syringoma | 103 (1.0%) | 103 (1.0%) | 16 (1.2%) |
| Venous lake | 101 (1.0%) | 101 (1.0%) | 13 (1.0%) |
| Wart | 636 (6.2%) | 636 (6.2%) | 83 (6.3%) |
| Xanthelasma | 18 (0.2%) | 18 (0.2%) | 2 (0.2%) |

The Severance validation dataset came from the Severance Hospital, Department of Dermatology and comprised both 34 benign and 9 malignant tumors.
The distribution of malignant tumors by anatomic location of the Severance Dataset A is as follows : head and neck 819 cases (7.9%), trunk 162 cases (1.6%), leg 153 cases (1.5%), arm 88 cases (0.8%). The distribution of benign tumors by anatomic location is as follows : head and neck 4,266 cases (40.9%), trunk 2,523 cases (24.2%), leg 1,433 cases (13.7%), arm 982 cases (9.4%).

### THE ALGORITHM

To validate the generalizability, the algorithm created in the previous study was tested again without modification in this study. The algorithm was trained not only with hospital archives but also with the archives generated with assistance of region-based convolutional neural network (RCNN) to reduce false positives. As a result, the algorithm was trained with 1,106,886 image crops. Algorithm has two parts: (A) lesion detection part and (B) disease classifier part. The lesion detection part was trained with faster RCNN (backbone=VGG-16) and CNN (SE-ResNet-50). The disease-classifier part was trained with CNNs (SENet and SE-ResNeXt-50).

The web-DEMO of the algorithm has been available in public (http://rcnn.modelderm.com) to facilitate scientific communications. We used the malignancy output which was predefined as: malignancy output = (basal cell carcinoma output + squamous cell carcinoma output + intraepithelial carcinoma output + keratoacanthoma output + malignant melanoma output) + 0.2 × (actinic keratosis output + ulcer output).

### PREDEFINED HIGH-SENSITIVITY THRESHOLD AND HIGH-SPECIFICITY THRESHOLD

We used two predefined thresholds (high-sensitivity threshold which was formerly named as T_90_ in the previous study and high-specificity threshold which was named as T_80_. These cut-off thresholds were defined as 90% or 80% sensitivity points with the dataset from Asan Medical Center, Department of Dermatology. In the dataset, all patients suspected of 10 major tumorous disorders (the same tumors as those of the Edinburgh dataset) from January 1, 2018 to June 30, 2018 were serially included. After pathologic confirmation, the dataset comprised of malignant tumors (81 patients), benign tumors (251 patients), and other various benign disorders (54 patients).

### UNPROCESSED IMAGE ANALYSIS VERSUS CROPPED IMAGE ANALYSIS

There are two different ways in using the algorithm for lesion diagnosis. The user or patient may localize the lesion of interest (cropped image analysis). If not, the algorithm needs to analyze unprocessed photograph without information on the lesion of interest (unprocessed image analysis). In unprocessed image analysis, our algorithm detected the lesion of interest in the photograph, and the highest malignancy output was used as the final output score among the malignancy outputs of suspected lesions. In cropped image analysis, the average malignancy output of the multiple lesions of interest was used as the final output score.

### READER TEST

A reader test was performed with diagnosis being made using only the lesions of interest without clinical information. After random sampling and shuffling of image sets of 1,320 patients, 44 test sets each containing 30 consecutive patients were created (Severance Dataset C; 44×30=1,320). A total of 44 dermatologists (5.7±5.2 years experiences after board-certification) received the different test sets, examined the multiple images of each patient, and chose their answer from one of 32 choices (=32 disorders for the multi-class classification; Table 3).

**Table 2.** Sensitivity, Specificity, Positive Predictive Value, and Negative Predictive Value of the Algorithm at Two Predefined Cutoff Thresholds.

| Disease | Classifier | Sensitivity<br>(95% CI) | Specificity<br>(95% CI) | PPV (95% CI) | NPV (95% CI) |
| --- | --- | --- | --- | --- | --- |
| All malignant tumors | Algorithm |  |  |  |  |
| 1,222 patients | High-Sensitivity Threshold | 79.1 (76.9–81.4) | 76.9 (76.1–77.8) | 31.3 (30.3–32.3) | 96.5 (96.2–96.9) |
|  | High-Specificity Threshold | 62.7 (59.9–65.5) | 90.0 (89.4–90.6) | 45.4 (43.7–47.3) | 94.8 (94.4–95.2) |
|  | Clinical Diagnosis |  |  |  |  |
|  | Top-1 | 70.2 | 95.6 | 68.1 | 96.0 |
|  | Top-2 | 84.9 | 86.0 | 44.6 | 97.7 |
|  | Top-3 | 88.1 | 83.8 | 41.9 | 98.1 |
| Basal cell carcinoma | Algorithm |  |  |  |  |
| 643 patients | High-Sensitivity Threshold | 81.3 (78.2–84.3) | 76.9 (76.0–77.8) | 19.8 (18.9–20.6) | 98.3 (98.0–98.6) |
|  | High-Specificity Threshold | 66.6 (63.0–70.1) | 90.0 (89.3–90.6) | 31.7 (30.0–33.6) | 97.5 (97.2–97.7) |
|  | Clinical Diagnosis |  |  |  |  |
|  | Top-1 | 74.0 | 95.6 | 54.2 | 98.1 |
|  | Top-2 | 87.7 | 86.0 | 30.4 | 99.0 |
|  | Top-3 | 90.4 | 83.8 | 28.0 | 99.2 |
| Squamous cell carcinoma | Algorithm |  |  |  |  |
| 158 patients | High-Sensitivity Threshold | 84.2 (78.5–89.9) | 76.9 (76.1–77.8) | 5.9 (5.5–6.3) | 99.7 (99.5–99.8) |
|  | High-Specificity Threshold | 70.9 (63.3–77.8) | 90.0 (89.4–90.6) | 10.8 (9.7–12.0) | 99.5 (99.3–99.6) |
|  | Clinical Diagnosis |  |  |  |  |
|  | Top-1 | 65.8 | 95.6 | 20.6 | 99.4 |
|  | Top-2 | 84.2 | 86.0 | 9.3 | 99.7 |
|  | Top-3 | 86.1 | 83.8 | 8.3 | 99.7 |
| Malignant melanoma | Algorithm |  |  |  |  |
| 83 patients | High-Sensitivity Threshold | 81.9 (73.5–90.4) | 76.9 (76.0–77.8) | 3.1 (2.8–3.4) | 99.8 (99.7–99.9) |
|  | High-Specificity Threshold | 61.4 (50.6–72.3) | 90.0 (89.4–90.6) | 5.3 (4.3–6.1) | 99.6 (99.5–99.7) |
|  | Clinical Diagnosis |  |  |  |  |
|  | Top-1 | 68.7 | 95.6 | 12.4 | 99.7 |
|  | Top-2 | 84.3 | 86.0 | 5.1 | 99.8 |
|  | Top-3 | 89.2 | 83.8 | 4.7 | 99.9 |

Algorithm analyzed unprocessed images from 1,222 patients of malignant tumor and 9,204 patients of benign tumor (Severance Dataset A). The result of each malignant tumor was calculated with the patients with malignant tumor of interest and the 9,204 patients with benign tumor.

**Table 3.** Multiclass Task – AUC and Top Accuracies of the Algorithm compared with Human Experts with the 32 Skin Tumors of the Severance dataset.

| No. | Class Name | Image Number | Clinical Diagnosis |  |  | Algorithm |  |  | AUC (95% CI) |
| --- | --- | --- | --- | --- | --- | --- | --- | --- | --- |
|  |  |  | Top-1 Accuracy | Top-2 Accuracy | Top-3 Accuracy | Top-1 Accuracy | Top-2 Accuracy | Top-3 Accuracy |  |
| 1 | Actinic keratosis | 784 | 68.8% | 78.4% | 80.2% | 26.4% | 42.1% | 52.3% | 0.925 (0.915–0.934) |
| 2 | Angiokeratoma | 39 | 66.7% | 69.2% | 69.2% | 46.2% | 56.4% | 64.1% | 0.931 (0.891–0.968) |
| 3 | Basal cell carcinoma | 643 | 64.5% | 75.0% | 77.3% | 47.0% | 59.7% | 66.6% | 0.915 (0.902–0.927) |
| 4 | Becker nevus | 14 | 85.7% | 85.7% | 85.7% | 42.9% | 42.9% | 50.0% | 0.994 (0.982–1.000) |
| 5 | Blue nevus | 115 | 71.3% | 78.3% | 79.1% | 71.3% | 82.6% | 84.3% | 0.967 (0.948–0.981) |
| 6 | Congenital nevus | 47 | 68.1% | 70.2% | 70.2% | 31.9% | 57.4% | 68.1% | 0.940 (0.891–0.975) |
| 7 | Dermatofibroma | 845 | 72.7% | 78.9% | 79.6% | 71.6% | 81.9% | 86.3% | 0.950 (0.942–0.958) |
| 8 | Epidermal cyst | 1501 | 80.7% | 86.2% | 87.1% | 56.0% | 70.2% | 77.5% | 0.956 (0.951–0.961) |
| 9 | Epidermal nevus | 26 | 53.8% | 69.2% | 69.2% | 26.9% | 26.9% | 30.8% | 0.907 (0.835–0.965) |
| 10 | Hemangioma | 263 | 43.0% | 50.6% | 52.1% | 30.8% | 46.8% | 57.4% | 0.875 (0.852–0.898) |
| 11 | Intraepithelial carcinoma | 255 | 43.1% | 51.8% | 54.9% | 16.5% | 28.6% | 34.5% | 0.888 (0.869–0.905) |
| 12 | Keratoacanthoma | 15 | 66.7% | 80.0% | 80.0% | 46.7% | 80.0% | 86.7% | 0.990 (0.978–0.997) |
| 13 | Lentigo | 67 | 43.3% | 55.2% | 55.2% | 35.8% | 47.8% | 53.7% | 0.870 (0.825–0.909) |
| 14 | Lymphangioma | 15 | 13.3% | 20.0% | 20.0% | 0.0% | 0.0% | 0.0% | 0.676 (0.550–0.798) |
| 15 | Malignant melanoma | 83 | 61.4% | 77.1% | 80.7% | 39.8% | 62.7% | 66.3% | 0.918 (0.888–0.945) |
| 16 | Melanocytic nevus | 1441 | 71.3% | 78.0% | 78.5% | 60.7% | 76.3% | 84.6% | 0.914 (0.906–0.922) |
| 17 | Mucocele | 73 | 84.9% | 87.7% | 89.0% | 27.4% | 45.2% | 47.9% | 0.985 (0.971–0.994) |
| 18 | Mucosal melanotic macule | 36 | 69.4% | 72.2% | 72.2% | 77.8% | 86.1% | 88.9% | 0.997 (0.995–0.999) |
| 19 | Neurofibroma | 199 | 51.8% | 57.3% | 57.8% | 43.7% | 57.3% | 62.3% | 0.943 (0.925–0.957) |
| 20 | Orgarnoid nevus | 62 | 80.6% | 88.7% | 88.7% | 62.9% | 79.0% | 82.3% | 0.966 (0.930–0.989) |
| 21 | Ota nevus | 24 | 79.2% | 83.3% | 83.3% | 54.2% | 66.7% | 75.0% | 0.999 (0.998–1.000) |
| 22 | Porokeratosis | 71 | 77.5% | 91.5% | 91.5% | 62.0% | 73.2% | 77.5% | 0.967 (0.942–0.987) |
| 23 | Poroma | 64 | 31.3% | 40.6% | 40.6% | 4.7% | 7.8% | 9.4% | 0.842 (0.787–0.889) |
| 24 | Portwine stain | 15 | 66.7% | 80.0% | 80.0% | 53.3% | 60.0% | 60.0% | 0.907 (0.780–0.998) |
| 25 | Pyogenic granuloma | 162 | 87.0% | 92.0% | 92.6% | 65.4% | 81.5% | 88.9% | 0.966 (0.945–0.981) |
| 26 | Seborrheic keratosis | 2370 | 65.1% | 78.4% | 79.3% | 48.5% | 64.6% | 72.2% | 0.910 (0.904–0.916) |
| 27 | Skin tag | 70 | 78.6% | 87.1% | 87.1% | 14.3% | 35.7% | 42.9% | 0.962 (0.945–0.977) |
| 28 | Squamous cell carcinoma | 158 | 44.9% | 69.6% | 72.2% | 28.5% | 47.5% | 56.3% | 0.912 (0.889–0.933) |
| 29 | Syringoma | 103 | 56.3% | 62.1% | 63.1% | 6.8% | 17.5% | 27.2% | 0.963 (0.942–0.981) |
| 30 | Venous lake | 101 | 81.2% | 86.1% | 87.1% | 40.6% | 55.4% | 65.3% | 0.980 (0.961–0.993) |
| 31 | Wart | 636 | 68.9% | 83.0% | 85.2% | 49.4% | 66.2% | 74.4% | 0.886 (0.875–0.898) |
| 32 | Xanthelasma | 18 | 94.4% | 100.0% | 100.0% | 72.2% | 88.9% | 88.9% | 0.998 (0.995–1.000) |
| Average |  | 322±541 | 65.4±17.7% | 73.9±16.6% | 74.7±16.6% | 42.6±20.7% | 56.1±22.8% | 61.9±22.9% | 0.931±0.062 |

Algorithm analyzed the Severance Dataset B (39,721 images of 10,315 patients; 32 disorders).
The images analyzed were cropped around lesion of interest.
We calculated the AUC value of ROC curve in a one-versus-rest manner.

### STATISTICAL ANALYSIS

We evaluated the performance of algorithm for the determination of malignancy. For the binary classification test, we draw ROC curve using the final output scores(the highest malignancy output in unprocessed image analysis and the average malignancy output in cropped image analysis ) and calculated area under the curve (pROC package version, 1.15.3; R version 3.4.4). The confidence interval was computed with 2,000 stratified bootstrap replicates.

## Results

### BINARY CLASSIFICATION

With the Dataset A, the algorithm analyzed all photographs of each patient for determining malignancy. The AUC value for determining malignancy was 0.863(95% CI 0.852–0.875).

At predefined high-sensitivity threshold, the sensitivity/specificity was 79.1%(76.9–81.4), 76.9%(76.1–77.8), and at predefined high-specificity threshold, the sensitivity/specificity was 62.7%(59.9–65.5), 90.0%(89.4–90.6) (Table 2). The sensitivity/specificity of the clinical diagnosis for malignancy determination calculated by clinical impressions were 70.2%/95.6%(Top1) and 88.1%/83.8%(Top3) (Figure 2). The differences between Top-1 clinical diagnosis and the algorithm at high-specificity threshold and the difference between Top-3 clinical diagnosis and the algorithm at high-sensitivity were statistically significant (McNemar test; all P<0.0001).

**Figures 2.**
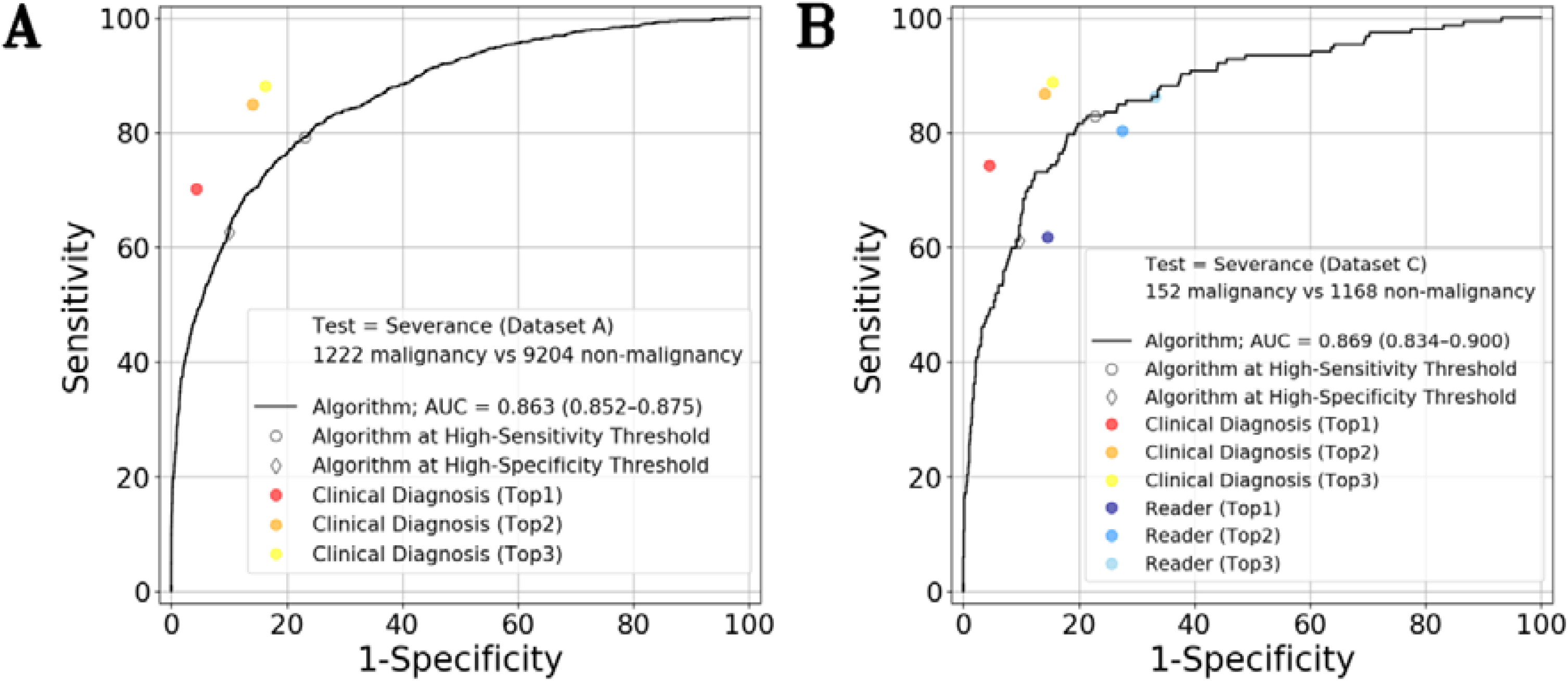
Diagnostic Performance for the Binary-Class Task of Determining Malignancy. (A) Test Dataset = Severance Dataset A (10,426 patients) (B) Test Dataset = Severance Dataset C (1,320 patients) Black curve – algorithm (unprocessed-images analysis) Diamond – algorithm at the predefined high-sensitivity Round – algorithm at the predefined high-specificity threshold Red, orange, and yellow circles – malignancy determination derived from Top-1, Top-2, and Top-3 clinical diagnoses of the 65 attending physicians Dark Blue, blue, and sky blue circles – malignancy determination derived from Top-1, Top-2, and Top-3 diagnoses of the 44 participants The Top-3 diagnosis corresponds to the algorithm at the high-sensitivity threshold, while the Top-1 diagnosis corresponds to the algorithm at the high-specificity threshold. In the reader test, the sensitivity / specificity of the algorithm at the predefined thresholds were 85.3±19.8% / 75.2±13.4% (high-sensitivity threshold) and 66.9±30.2% / 87.4±16.5% (high-specificity threshold). The sensitivity / specificity of the participants were 84.9±24.8% / 66.9±15.1% (Top-3) and 65.8±33.3% / 85.7±11.0% (Top-1). The sensitivity/specificity of the clinical diagnoses were 90.6±15.5% / 84.6±7.1% (Top-3) and 76.8±20.5% / 95.6±4.2% (Top-1). Overall, the performance of the clinical diagnoses were superior to those of both the algorithm and participants to the reader test. The performance between the algorithm and participants was comparable.

The positive predictive value (PPV) of the algorithm were 31.3%(30.3–32.3) at high-sensitivity threshold and 45.4%(43.7–47.3) at high-specificity threshold, and the PPVs of Top-1 and 3 clinical diagnosis were 68.1%, 41.9%. The negative predictive value (NPV) of the algorithm were 96.5%(96.2–96.9) at high-sensitivity threshold and 94.8%(94.4–95.2) at high-specificity threshold, and the NPVs of Top-1 and 3 clinical diagnosis were 96.0% and 98.1%.

The performances of two analysis methods (unprocessed image analysis versus cropped image analysis) were compared with 39,721 images from 10,315 patients (Figure 3). The AUC of 0.881(0.870–0.891) in cropped image analysis was slightly better than the AUC of 0.870(0.858-0.881) in unprocessed image analysis (DeLong’s test; P=0.0022).

**Figures 3.**
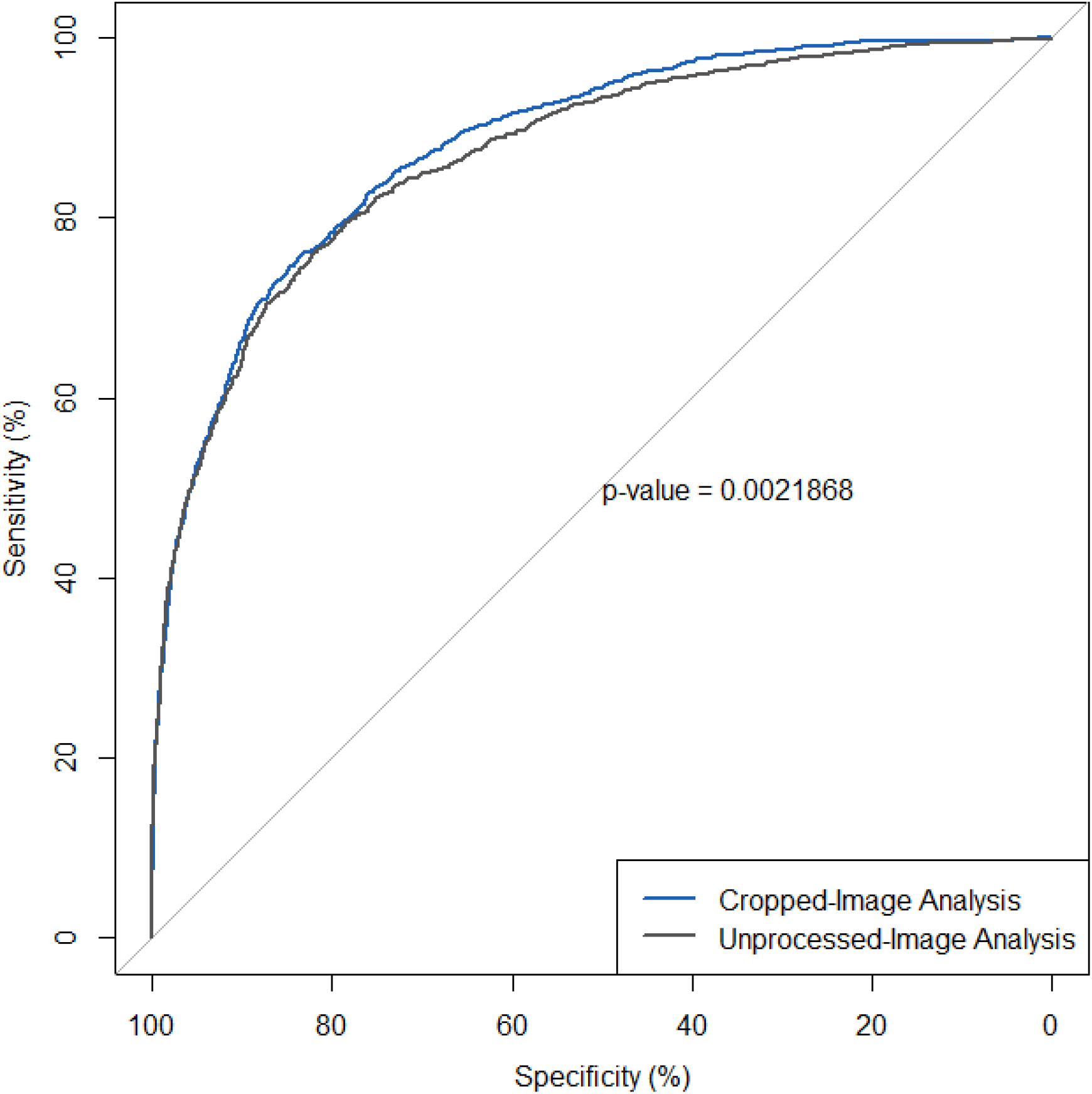
Performance Comparison Between Unprocessed Image Analysis and Cropped Images Analysis. Black curve – algorithm (unprocessed-image analysis) Gray curve – algorithm (cropped-image analysis) Severance Dataset B (39,721 images from 10,315 patients) was used for the comparison. As shown in Figure 1, the images of small classes and untrained classes were not used for this test. In cropped-image analysis, algorithm analyzed cropped lesion of interest which was pointed out by user. In unprocessed-image analysis, algorithm detected location of lesion and analyzed the suspected lesions. The AUC in cropped-images analysis were 0.881 (95% CI 0.870–0.891), which was slightly higher than the AUC of 0.870 (95% CI 0.858–0.881) in unprocessed-image analysis.

### MULTICLASS CLASSIFICATION WITH 32 SKIN TUMORS OF THE SEVERANCE DATASET

For multi-class classification test, we used AUC and Top-accuracy as evaluation metrics. We calculated both Top accuracy of total patients and Top accuracy of each classes because the number of patients varies in each classes.

The calculation of Top accuracy of each classes was performed with 32 disorders (Severance Dataset B; 39,721 images from 10,315 patients). We excluded 6 classes (angiofibroma, Café au lait macule, juvenile xanthogranuloma, milia, nevus spilus, and sebaceous hyperplasia) of which the number of patient was less than 10, and we also excluded 5 classes (Spiz nevus, dermatofibrosarcoma protuberans, angiosarcoma, Kaposi’s sarcoma, and Merkel cell carcinoma) which were not trained to the algorithm. We calculated mean accuracy by averaging the accuracies of 32 disorders as follows: macro-averaged mean Top-(n) Accuracy = (Top-(n) Accuracy of actinic keratosis + Top-(n) Accuracy of angiokeratoma + … + Top-(n) Accuracy of xanthelasma) / 32. The macro-averaged mean Top-1 and 3 accuracies of clinical diagnosis were 65.4±17.7% and 74.7±16.6%, and those of the algorithm were 42.6±20.7% and 61.9±22.9%, respectively. In order to reflect the difference in the number of cases for each disease, the micro-averaged accuracy was calculated as follows: micro-averaged mean Top-(n) Accuracy = (Top-(n) matched cases in total) / 10,315. The micro-averaged mean Top-1 and 3 accuracies of clinical diagnosis were 68.2% and 78.7% and those of the algorithm were 49.2% and 71.2%, respectively.

The mean AUC of 32 classes was 0.931±0.062 (Figure 4 and Table 3).

**Figure 4.**
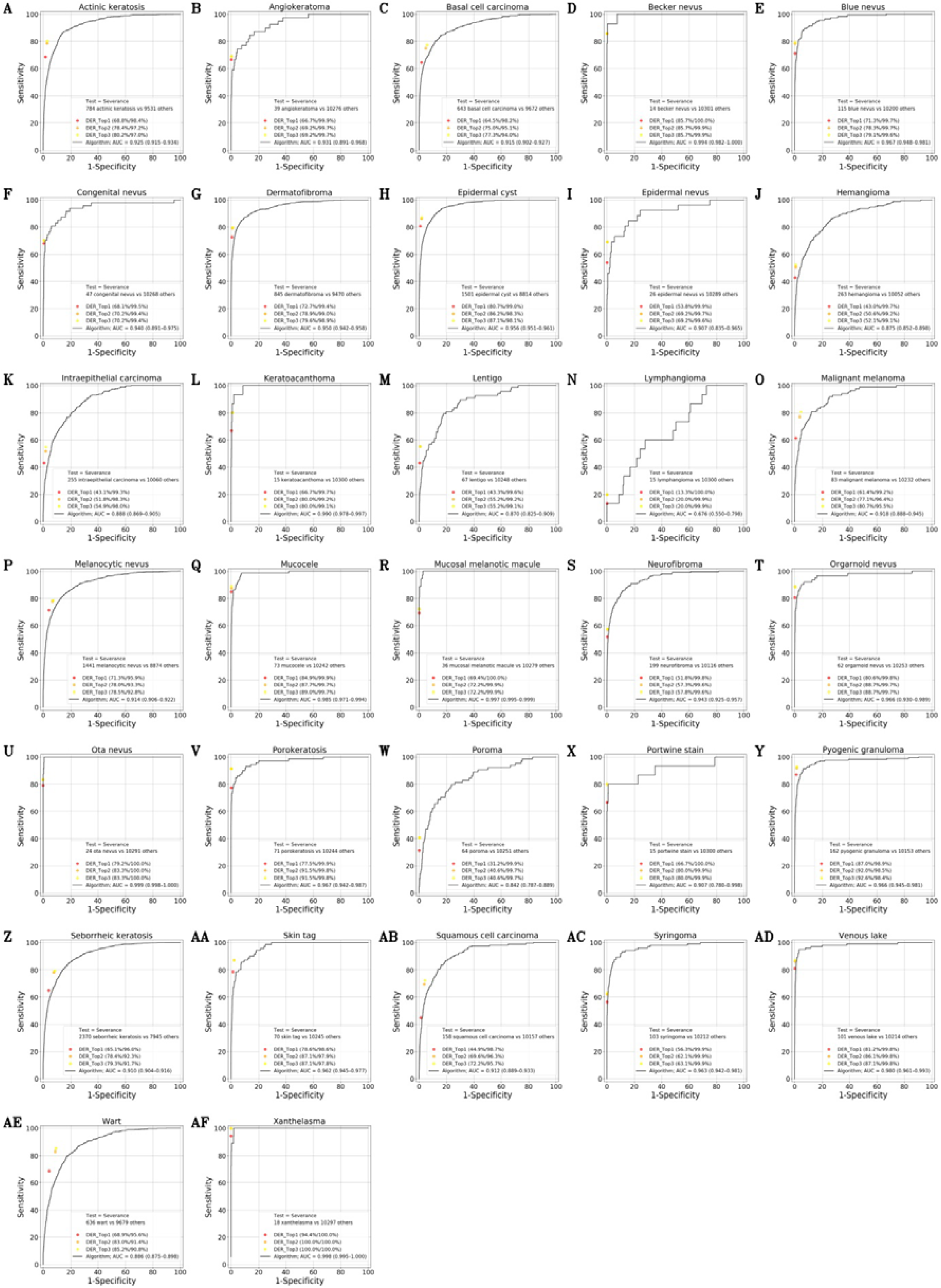
Multiclass Task – The ROC of the Algorithm with 32 Skin Tumors of the Severance dataset. Algorithm analyzed the Severance Dataset B (39,721 images of 10,315 patients; 32 disorders). The images analyzed were cropped around lesion of interest.

### MULTICLASS CLASSIFICATION WITH 10 SKIN TUMORS OF THE EDINBURGH DATASET

We performed an additional external validation with the Edinburgh dataset. The Edinburgh dataset consists mainly of Caucasians and is commercially available dataset with 10 benign and malignant skin tumor (Table 4; https://licensing.edinburgh-innovations.ed.ac.uk/i/software/dermofit-image-library.html). In previous studies^4,5^, the Edinburgh dataset was used as a validation dataset.

**Table 4.**
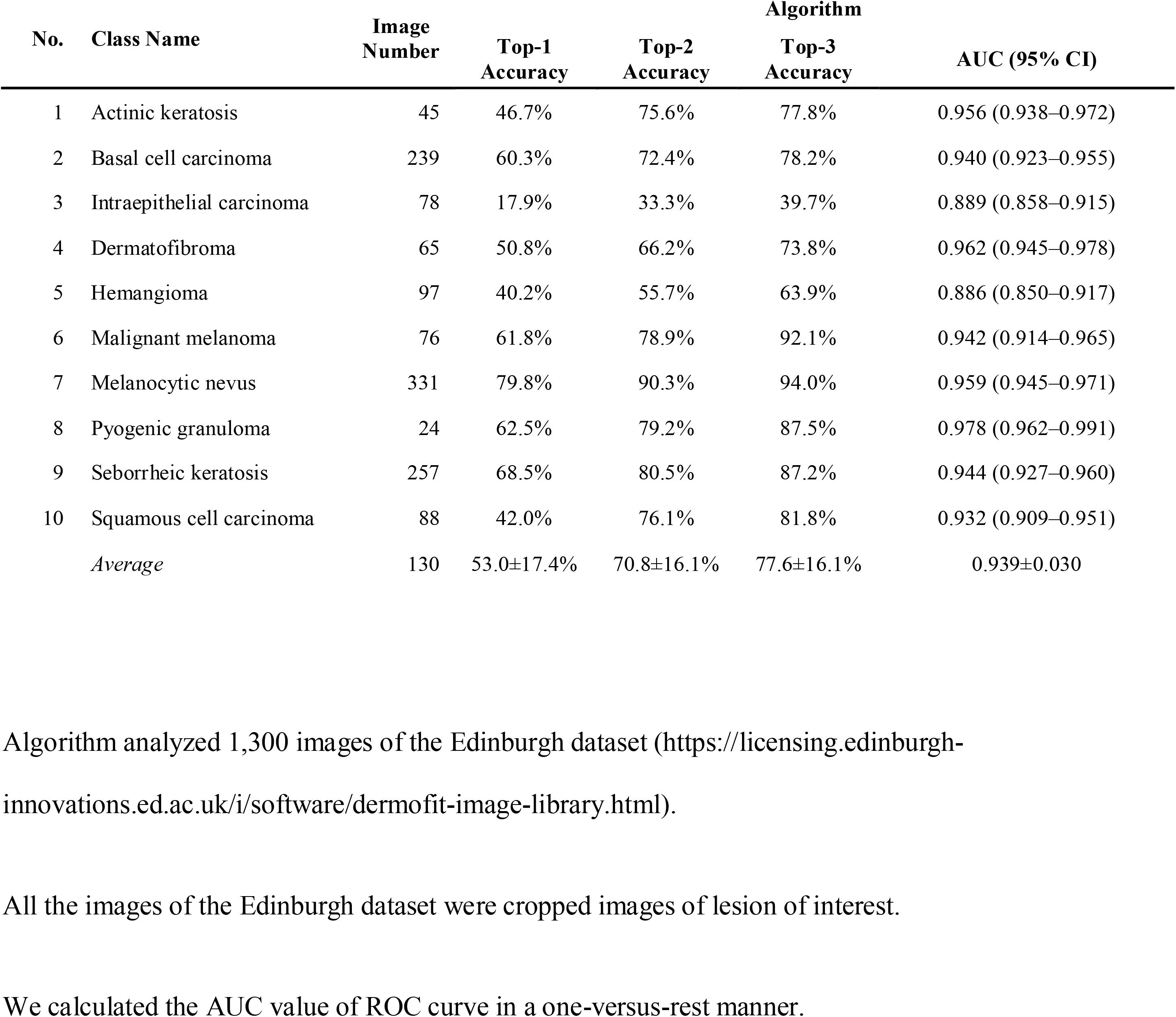
Multiclass Task – AUC and Top Accuracies of the Algorithm with 10 Skin Tumors of the Edinburgh dataset.

For the multi-class classification, the macro-averaged mean Top-1 and 3 accuracies were 53.0% and 77.6%, respectively and the mean AUC of each classes was 0.939±0.030 (Figure 5 and Table 4).

**Figure 5.**
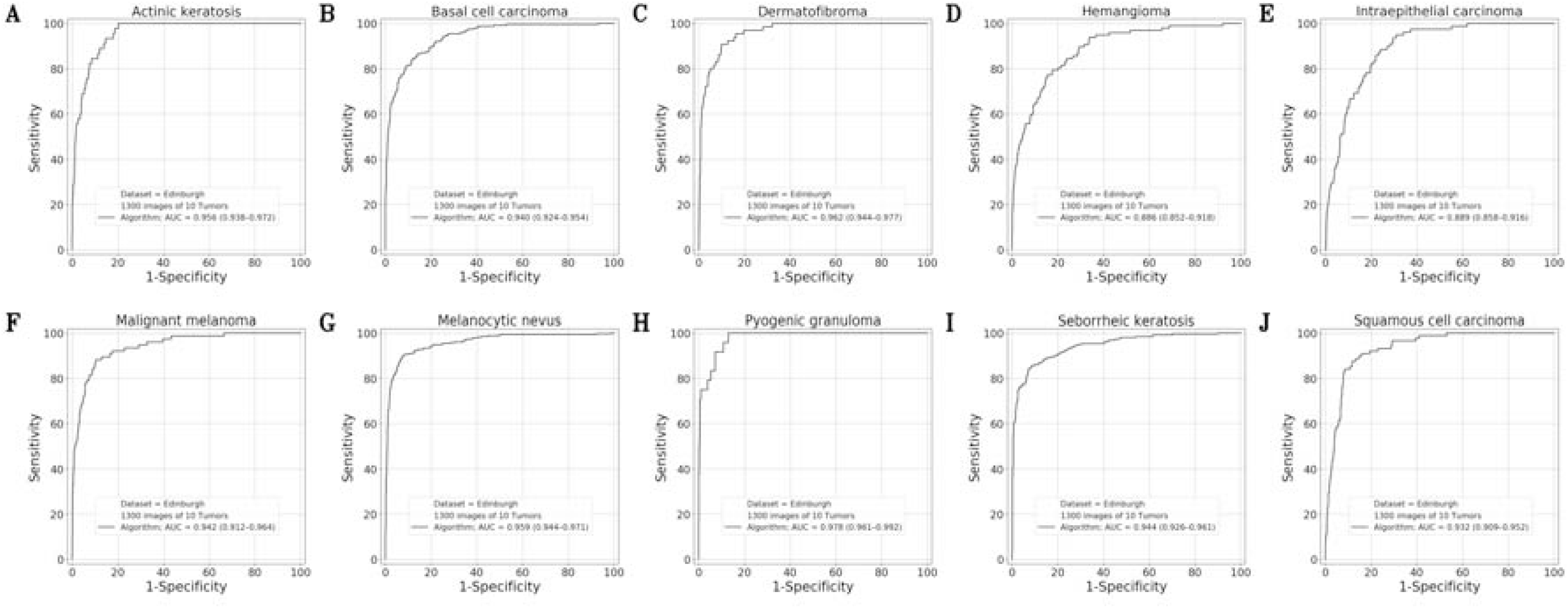
Multiclass Task – The ROC curve of the Algorithm with 10 Skin Tumors of the Edinburgh dataset. Algorithm analyzed 1,300 images of the Edinburgh dataset (https://licensing.edinburgh-innovations.ed.ac.uk/i/software/dermofit-image-library.html). All the images of the Edinburgh dataset were cropped images of lesion of interest. We draw the ROC curve in a one-versus-rest manner.

### READER TEST WITH BOARD-CERTIFIED DERMATOLOGISTS

A reader test was conducted with each of 44 dermatologists evaluating a different test set including images from 30 randomly selected patients (Severance Dataset C).

For the binary classification of determining malignancy, we compared the diagnoses of the participants in the reader test and those of the algorithm. As shown in Figure 2B, overall, the sensitivities and specificities of the algorithm were comparable with those of the participants (Wilcoxon test; P=0.99/0.0043 for sensitivity/specificity of the Top-3 of the participants versus sensitivity/specificity of the algorithm at the high-sensitivity threshold; P=0.61/0.097 for sensitivity/specificity of the Top-1 of the participants versus sensitivity/specificity of the algorithm at the high-specificity threshold).

For the multi-class classification to derive the exact diagnoses, the Top-1 / Top-3 accuracies of the algorithm were 49.5%±9.8% / 69.4%±7.5%, and those of the participants were 37.7%±10.9% / 48.3%±11.5%, and those of the clinical diagnoses were 68.1%±9.9% / 77.3%±9.6%. The Top accuracies of the algorithm were superior to those of the participants (Wilcoxon test; all Top 1~3 accuracies; P<0.0001), whereas the Top accuracies of the algorithm were inferior to those of the clinical diagnosis (Wilcoxon test; all Top 1~3 accuracies; P<0.0001).

## Discussion

Deep learning algorithm has recently shown remarkable performance in dermatology. With clinical photographs^4-8^ and dermoscopic images^4,7,10-13^, algorithm showed comparable or superior performance to dermatologists with regards to the problem of malignant melanoma vs nevus, and carcinoma vs benign keratotic tumors. Direct comparison between the results of various studies is difficult because inclusion and exclusion criteria of validation dataset are unclear and the validation datasets were private^18^. To date, there have been only few studies that compared the algorithm’s performance with experts using external datasets.^19^ It was impossible to test their algorithm in most studies and it was unknown whether the algorithm could reproduce similar performance with other datasets. In fact, several studies report that currently available smartphone applications did not show the dermatologist-level performance as shown in academic papers.^20,21^

When investigating the performance of deep learning algorithm with skin diseases, there are various factors that result in poor reproducibility or generalizability in real practice.

1. Limited number of training classes: the number of disorders in validation dataset is usually much smaller than that in real practice^17^. Algorithm had shown good performance with a narrow range problem such as “melanoma vs nevus”. However, as shown in Table 4, the mean Top-1 accuracy of clinical diagnosis was only 65.4%, which suggests that it is hard to choose the initial problem correctly. For example, the algorithm trained only with melanoma and nevus may be misused for the diagnosis of other disorder such as seborrheic keratosis, which result in epistemic uncertainty.^16^
2. Innocent biases of internal validation: in internal validation, prediction may be drawn from unexpected false features instead of true features of disease. The composition of image and the presence of skin markings were likely to affect the prediction of algorithm.^22,23^ From the perspective that algorithm may be trained with unexpected structured noise, the current convolutional neural network draws a statistical prediction from massive image data, rather than real intelligence.
3. Preselection of specialist: Preselection bias exists if the image is acquired in real time procedure, such as endoscopy, ultrasonography, and clinical photography. In dermatology, both training and test datasets are created by dermatologist and dermatologist determines which lesions need to be included and which composition is adequate. Because algorithm shows aleatory uncertainty, we may not get the same performance without the preselection of dermatologist.
4. Unclear inclusion and exclusion criteria: selection bias affects the result if the cases with negative results were excluded more frequently for any reason. All patients in the archives should be selected based on specific exclusion criteria to prevent selection bias.
5. Specific optimization: hyperparameters of algorithm may be specifically optimized to the validation dataset at the test time, and lack of generalizability for other dataset.
6. Data leakage: data leakage occurs when training and test dataset were not split completely, or when the data set framework is designed incorrectly. In case of data leakage, model uses inessential additional information for classification.^24^
7. Non-representativeness of training dataset^25^: hospital achieve include extraordinary case which requires biopsy. For example, there are many cases of small melanoma which does not have characteristic bizarre morphology. If algorithm was trained through these extraordinary cases, it may show many false positives. In addition, hospital achieve does not usually have enough number of images of common disorders, which reduces the diagnostic accuracy for common disorders.
8. Data relevancy: although algorithm analyze image better than dermatologist, dermatologist in real practice shows much better diagnostic accuracy because dermatologist considers all patient’s information for diagnosis.

To minimize biases, this study was designed with the following considerations. 1) To prevent uncertainty for untrained classes, we included almost all kinds of skin tumors (43 disorders). 2) To avoid selection bias, all biopsied cases in one hospital were included with exclusion criteria. 3) In order to prevent innocent bias from data leakage, we tested with two external validation datasets, the Severance dataset and Edinburgh dataset. 4) To show generalizability, we verified previously created algorithm with new validation dataset built later. 5) The diagnostic accuracy of algorithm was compared with that of attending physician who considered all information of patient for diagnosis. 6) Algorithm was tested with unprocessed images without preselection of dermatologist.

To date, most comparisons between dermatologist and algorithm have been performed using single cropped image of lesions. Deep learning algorithm has shown comparable performance with dermatologists in the same settings.^4,5,7,8,11-13^ Meta-analysis of medical studies also showed that, under the same conditions, the performance of algorithms was comparable to that of healthcare specialists.^19^

In this study, we investigated the diagnostic accuracy of the dermatologist in real practice with biopsied cases. Skin cancer diagnosis is a difficult task for experienced dermatologists even when given all clinical information. The sensitivity which calculated from three differential impressions of the dermatologists was 88.1%, meaning that 11.9% of malignant tumors were ignored by the dermatologists even after thorough examination, but the presence of malignancy was notified later after receiving the biopsy report. The malignancy prediction derived from the first clinical impression was 70.2%, implying that there was 29.8% misdiagnosis in the initial impression. In the multiclass classification which requires correct diagnosis, the mean Top-1 and 3 accuracy of the clinical diagnosis were lower as 65.4±17.7% and 74.7±16.6% as shown in Table 4. Deep learning algorithm shows uncertainty for untrained task^16^, therefore, if the algorithm is not an unified classifier but a classifier trained with limited classes, the precondition may be wrong^26^.

The performance of attending physician significantly improves after considering various information of patient. In the actual medical practice, physician consider not only the visual information of the lesion but also various information such as previous medical history, referral note, demographics, family history, and physical examination, to improve the diagnostic accuracy. In radiology or pathology, the actual reader does not diagnose with image alone, but they also checks medical history through chart review and reflects clinicopathologic correlation. At present, there has been several reports^27-29^ to reflect patient’s metadata with multimodal approach. The results obtained by combining images and patient clinical information showed a general improvement of around 7% in accuracy.^27^ In this study, the Top-1 accuracy of the clinical diagnoses (68.1±9.9%) was much better than that of the 44 dermatologists (37.7±10.9%) in the reader test with the Severance C dataset.

Algorithms may play an important role in the screening of suspected patients if mass diagnostics task will be done only with image information. Our algorithm showed a comparable performance in full automatic mode regardless of preselection of the lesion (AUC with unprocessed images = 0.870 vs AUC with cropped image = 0.881). In previous study^9^ with unprocessed facial images, our algorithm showed an AUC of 0.896 with the internal validation (386 patients; 81 with malignant tumor and 305 with benign tumor) and the performance of algorithm was comparable with that of 13 dermatologists in terms of F1 score and Youden index. We reproduced a similar AUC of 0.863 with large scale external validation dataset.

Algorithm can work endlessly with minimal cost. In case of cancer screening, clinical photographs may be available alone without other information. In this study, the sensitivity at the high-sensitivity threshold was 79.1%, which was located between the sensitivities (70.2% and 84.9%) derived from the Top-2 and Top-1 clinical diagnoses. In the previous study, approximately 50% images of malignant cases could be misinterpreted to benign by general public^9^. We expect algorithm-based cancer screening helps appropriate referral to dermatology in the future.

### LIMITATION

Although we obtained a mean AUC of 0.939±0.030 (Table 4) with the Edinburgh dataset (1,300 images of 10 benign and malignant tumors) which mainly consists of Caucasians, we need to test our algorithm with unprocessed clinical photographs of various race/ethnicity. With the identical image of dysplastic nevus, we should warn a high chance of melanoma for Caucasian, but may recommend close-observation for Asian because melanoma is a rare disorder in Asian. We also need to test with Asian in other regions because disease prevalence may vary from region to region even if the races are the same.

In this study, most of training and validation images were taken with adequate quality. The performance of algorithm could be affected by image quality more than that of human.^22,30^ Therefore, the performance of the algorithm on the photographs taken by the public must be verified in further study.

## CONCLUSION

In this study, the attending physician in real practice made more accurate diagnosis because not only image information but also clinical information was considered to make a diagnosis. Without preselection of lesion by dermatologist, our algorithm showed the AUC of 0.863, which implies unprecedented potential as a mass screening tool. More clinical information should be integrated for more accurate prediction of the algorithm in the future.

## ACKNOWLEDGMENTS

Thanks to PhD, Park Jeoongsung in Qualcomm for helping to stabilize the web-DEMO.

## AUTHOR CONTRIBUTIONS

Han and Moon designed experiments.

Han, Moon, Lee, Na, Kim MS, Park GH, and Kim SH prepared validation datasets.

Han, Moon, and Chang performed experiments.

Han, Moon, Park, Kim MS, Na, and Kim KW interpreted results.

Han, Moon, and Kim KW wrote the manuscript and prepared the figures.

Chang and Lee supervised the study.

Han created and managed the web-DEMO.

All authors approved the final version of the manuscript.

## DATA AVAILABILITY

The images used to test the neural networks described in the manuscript are subject to privacy regulations and cannot be made available in totality. The test subset may be available upon a reasonable request and an approval of IRB of the originating university hospital.

## Data Availability

**Abbreviations**
AUC: area under the curve;
CNN: convolutional neural network;
PPV: positive predictive value;
ROC curve: receiver operating characteristic curve;
RCNN: region-based convolutional neural network;
NPV: negative predictive value

## REFERENCES

1. Gulshan V, Peng L, Coram M, et al. Development and validation of a deep learning algorithm for detection of diabetic retinopathy in retinal fundus photographs. Jama 2016;316:2402–10.

2. Rajpurkar P, Irvin J, Ball RL, et al. Deep learning for chest radiograph diagnosis: A retrospective comparison of the CheXNeXt algorithm to practicing radiologists. PLoS medicine 2018;15:e1002686.

3. Chilamkurthy S, Ghosh R, Tanamala S, et al. Deep learning algorithms for detection of critical findings in head CT scans: a retrospective study. The Lancet 2018;392:2388–96.

4. Esteva A, Kuprel B, Novoa RA, et al. Dermatologist-level classification of skin cancer with deep neural networks. Nature 2017;542:115–8.

5. Han SS, Kim MS, Lim W, Park GH, Park I, Chang SE. Classification of the Clinical Images for Benign and Malignant Cutaneous Tumors Using a Deep Learning Algorithm. Journal of Investigative Dermatology 2018.

6. Fujisawa Y, Otomo Y, Ogata Y, et al. Deep learning-based, computer-aided classifier developed with a small dataset of clinical images surpasses board-certified dermatologists in skin tumor diagnosis. British Journal of Dermatology 2018.

7. Tschandl P, Rosendahl C, Akay BN, et al. Expert-Level Diagnosis of Nonpigmented Skin Cancer by Combined Convolutional Neural Networks. JAMA dermatology 2018.

8. Cho S, Sun S, Mun J, et al. Dermatologist-level classification of malignant lip diseases using a deep convolutional neural network. British Journal of Dermatology 2019.

9. Han SS, Moon I J, Lim W, et al. Keratinocytic Skin Cancer Detection on the Face Using Region-Based Convolutional Neural Network. JAMA dermatology 2019.

10. Haenssle H, Fink C, Schneiderbauer R, et al. Man against machine: diagnostic performance of a deep learning convolutional neural network for dermoscopic melanoma recognition in comparison to 58 dermatologists. Annals of Oncology 2018.

11. Brinker TJ, Hekler A, Enk AH, et al. A convolutional neural network trained with dermoscopic images performed on par with 145 dermatologists in a clinical melanoma image classification task. European Journal of Cancer 2019;111:148–54.

12. Maron RC, Weichenthal M, Utikal JS, et al. Systematic outperformance of 112 dermatologists in multiclass skin cancer image classification by convolutional neural networks. European Journal of Cancer 2019;119:57–65.

13. Tschandl P, Codella N, Akay BN, et al. Comparison of the accuracy of human readers versus machine-learning algorithms for pigmented skin lesion classification: an open, web-based, international, diagnostic study. The Lancet Oncology 2019.

14. Topol EJ. High-performance medicine: the convergence of human and artificial intelligence. Nature medicine 2019;25:44.

15. Silver D, Huang A, Maddison CJ, et al. Mastering the game of Go with deep neural networks and tree search. Nature 2016;529:484.

16. Kendall A, Gal Y. What uncertainties do we need in bayesian deep learning for computer vision? Advances in neural information processing systems; 2017. p. 5574-84.

17. Tschandl P. Problems and Potentials of Automated Object Detection for Skin Cancer Recognition. JAMA dermatology 2019.

18. Brinker TJ, Hekler A, Utikal JS, et al. Skin cancer classification using convolutional neural networks: systematic review. Journal of medical Internet research 2018;20:e11936.

19. Liu X, Faes L, Kale AU, et al. A comparison of deep learning performance against health-care professionals in detecting diseases from medical imaging: a systematic review and meta-analysis. The Lancet Digital Health 2019;1:e271-e97.

20. Chuchu N, Takwoingi Y, Dinnes J, et al. Cochrane Skin Cancer Diagnostic Test Accuracy Group. Smartphone applications for triaging adults with skin lesions that are suspicious for melanoma, status and date: New, published in 2018.

21. Xiong M, Pfau J, Young AT, Wei ML. Artificial Intelligence in Teledermatology. Current Dermatology Reports 2019;8:85–90.

22. Navarrete-Dechent C, Dusza SW, Liopyris K, Marghoob AA, Halpern AC, Marchetti MA. Automated Dermatological Diagnosis: Hype or Reality? Journal of Investigative Dermatology 2018.

23. Winkler JK, Fink C, Toberer F, et al. Association between surgical skin markings in dermoscopic images and diagnostic performance of a deep learning convolutional neural network for melanoma recognition. JAMA dermatology 2019.

24. Nisbet R, Elder J, Miner G. Handbook of statistical analysis and data mining applications: Academic Press; 2009.

25. Parikh RB, Teeple S, Navathe AS. Addressing Bias in Artificial Intelligence in Health Care. JAMA 2019.

26. Narla A, Kuprel B, Sarin K, Novoa R, Ko J. Automated classification of skin lesions: from pixels to practice. Journal of Investigative Dermatology 2018;138:2108–10.

27. Pacheco AG, Krohling RA. The impact of patient clinical information on automated skin cancer detection. arXiv preprint arXiv:190912912 2019.

28. Yap J, Yolland W, Tschandl P. Multimodal skin lesion classification using deep learning. Experimental dermatology 2018;27:1261–7.

29. Liu Y, Jain A, Eng C, et al. A deep learning system for differential diagnosis of skin diseases. arXiv preprint arXiv: 190905382 2019.

30. Dodge S, Karam L. A study and comparison of human and deep learning recognition performance under visual distortions. 2017 26th international conference on computer communication and networks (ICCCN); 2017: IEEE. p. 1–7.

